# Comparison of Antibody Response Durability of mRNA-1273, BNT162b2, and Ad26.COV2.S SARS-CoV-2 Vaccines in Healthcare Workers

**DOI:** 10.1101/2022.01.14.22269297

**Authors:** Wendy M. Brunner, Daniel Freilich, Jennifer Victory, Nicole Krupa, Melissa B. Scribani, Paul Jenkins, Emily G. Lasher, Amanda Fink, Anshini Shah, Peggy Cross, Valerie Bush, Laura J. Peek, Gary A. Pestano, Anne M. Gadomski

## Abstract

**Importance:** There is a dearth of comparative immunologic durability data after COVID-19 vaccinations.

**Objective:** To compare antibody responses and vaccine effectiveness 8.4 months post-primary COVID-19 vaccination.

**Design:** Setting and Participants: In this cohort study of 903 healthcare workers who completed surveys about baseline characteristics and COVID-19 vaccine/infection history, 647 had antibody assays completed and were included herein.

**Exposure:** COVID-19 vaccination with mRNA-1273 (n=387); BNT162b2 (n=212); or Ad26.COV2.S (n=10); unvaccinated (n=10); or boosted (n=28).

**Main Outcomes and Measures:** The primary outcome was IgG anti-spike titer. Secondary/tertiary outcomes included IgG spike receptor-binding domain competitive antibody blocking ELISA pseudoneutralization against the USA-WA1/2020 strain, and vaccine effectiveness against COVID-19 infection. Antibody levels were compared using ANOVA and multiple linear regression.

**Results:** Mean age was 49.7, 75.3% were female, and mean comorbidities/patient was 0.7. Baseline variables were balanced (p>.05) except for immunosuppression (higher in boosted, p=.047), prior COVID-19 infections (higher with Ad26.COV2.S and unvaccinated, p<.001), and time since primary vaccination (higher with mRNA-1273 and BNT162b2 than Ad26.COV2.S, p<.001).

Unadjusted median (IQR) IgG anti-spike titers (AU/mL) were 1539.5 (876.7-2626.7) for mRNA-1273, 751.2 (422.0-1381.5) for BNT162b2, 451.6 (103.0-2396.7) for Ad26.COV2.S, 113.4 (3.7-194.0) for unvaccinated, and 31898.8 (21347.1-45820.1) for boosted (mRNA-1273 vs. BNT162b2, p<.001; mRNA-1273, BNT162b2, or boosted vs. unvaccinated, p<.006; mRNA-1273, BNT162b2, Ad26.COV2.S, or unvaccinated vs. boosted, p<.001; all other comparisons, p>.05). Unadjusted median (IQR) pseudoneutralization percentages were 90.9% (80.1-95.0) for mRNA-1273, 77.2% (59.1-89.9) for BNT162b2, 57.9% (36.6-95.8) for Ad26.COV2.S, 40.1% (21.7-60.6) for unvaccinated, and 96.4% (96.1-96.6) for boosted (mRNA-1273 vs. BNT162b2, p<.001; mRNA-1273, BNT162b2, or boosted vs. unvaccinated, p<.028; mRNA-1273, BNT162b2, Ad26.COV2.S, or unvaccinated vs. boosted, p<.001; all other comparisons, p>.05). Adjusted anti-spike and pseudoneutralization comparisons of mRNA-1273 and BNT162b2 showed similar patterns (p<.001). Vaccine effectiveness was 87-89% for mRNA-1273, BNT162b2, and boosted, and 33% for Ad26.COV2.S; no group differences were statistically significant.

**Conclusions and Relevance:** Durability of antibody responses 8.4 months after COVID-19 primary vaccination was significantly higher with mRNA-1273 than with BNT162b2, however, vaccine effectiveness was equivalent. Antibody responses and vaccine effectiveness were lower but not significantly different for Ad26.COV2.S; given statistical uncertainty in the small Ad26.COV2.S group, clinically important effects cannot be excluded.

## Introduction

To date, more than 820,000 Americans have died of COVID-19. Vaccination with FDA approved/authorized COVID-19 vaccines is imperative to control the pandemic. A number of studies have shown robust similar or higher early anti-spike (anti-S), anti-S-receptor binding domain (RBD), and neutralizing antibody (nAb) responses with mRNA-1273 than BNT162b2 and higher responses with both mRNA vaccines than Ad26.COV2.S.^1-7^ The pattern for early vaccine effectiveness (VE) is similar, mRNA-1273 being about 90-95%, BNT162b2 about 90%, and Ad26.COV2.S about 70-85%, with lower and higher percentages being against infection and severe disease, respectively.^5, 8-11^

Individual durability studies for these vaccines have shown waning immunity over time, as measured by decreasing antibody titers and VE (breakthrough infections).^12-34^ The literature for comparative vaccine antibody durability, however, is limited. One study showed marked decreases in antibodies over 6-8 months for mRNA-1273 and BNT162b2 which contrasted with relatively durable responses for Ad26.COV2.S; because responses to Ad26.COV2.S were lower early post-vaccination, antibody responses were comparable at 6-8 months.^3^ Another comparative antibody durability study showed 3-fold decreases in anti-S and anti-S-RBD antibody titers from peak post-vaccination (14-119 days) to >120 days with mRNA-1273 and BNT162b2, but mRNA-1273 titers were 3-fold higher at both time points.^35^ Numerous comparative VE durability studies, occurring at different pandemic stages with differing prevalent strains, showed decreases to 80-95% at 3-7 months for mRNA-1273, 65-90% for BNT162b2, and 60-70% for Ad26.COV2.S.^5, 10, 11, 35-38^

Due to the dearth of comparative vaccine durability studies, particularly for antibody responses, we sought to compare medium range durability (at 8.4 months) of antibody responses in a cohort of healthcare workers (HCWs) in the Bassett Healthcare Network (NY, USA) vaccinated with FDA-approved/authorized COVID-19 vaccines.

Our primary objective was to compare medium range (median 8.4 months) SARS-CoV-2 anti-S antibody titers in HCWs vaccinated with mRNA-1273, BNT162b2, or Ad26.COV2.S, or unvaccinated, or vaccine-boosted. Our secondary objective was a similar comparison for IgG-S-RBD competitive antibody blocking ELISA pseudoneutralization semiquantitative inhibition percentages against the USA-WA1/2020 strain, at a median of 8.4 months. Tertiary objectives included: comparisons of COVID-19 infection rates based on immunological parameters (IgG nucleocapsid (anti-N) seroconversion from an early pandemic baseline)) and clinical parameters (clinical infections based on self-reported survey and PCR swab positivity) and VE percentages (against COVID-19 infection); comparisons of antibody responses stratified by age, number of comorbidities, immunosuppression, time since primary vaccination, and COVID-19 infection; and assessments of correlation between anti-S and pseudoneutralization results.

## Methods

### Design and Setting

This observational cohort study is a follow-up to a seroprevalence study conducted among Bassett Healthcare Network HCWs in May-August 2020 (IRB#1597947).^39^ Primary steps included securing consent, online surveys, blood drawing/processing, and antibody assays. The Mary Imogene Bassett IRB approved this follow-up as an amendment to the original study.

### Participants, Data Collection, and Interventions

HCWs were eligible for this follow-up study if they consented to storage of their plasma samples for future research and were available for blood drawing regardless of vaccination or infection status. Of 2056 HCWs, 1806 were invited to participate and 903 (50%) consented and completed the study’s survey. Among them, 653 (72%) had blood drawn September 23 – November 16, 2021; 238 did not have blood drawn because of scheduling/logistics challenges, 12 declined, and 6 were excluded because of incomplete primary vaccination, yielding 647 HCWs for inclusion (eFigure 1), stratified into 4 groups: (1) mRNA-1273-vaccinated; (2) BNT162b2-vaccinated; (3) Ad26.COV2.S-vaccinated; and (4) unvaccinated. A fifth group—boosted (regardless of vaccine type)—was added upon FDA/CDC recommendations for boosting of HCWs in October 2021. Primary vaccination was defined as two mRNA vaccine doses or one Ad26.COV2.S dose; “boosted” was defined as primary vaccination plus an additional vaccine.

**Figure 1.**
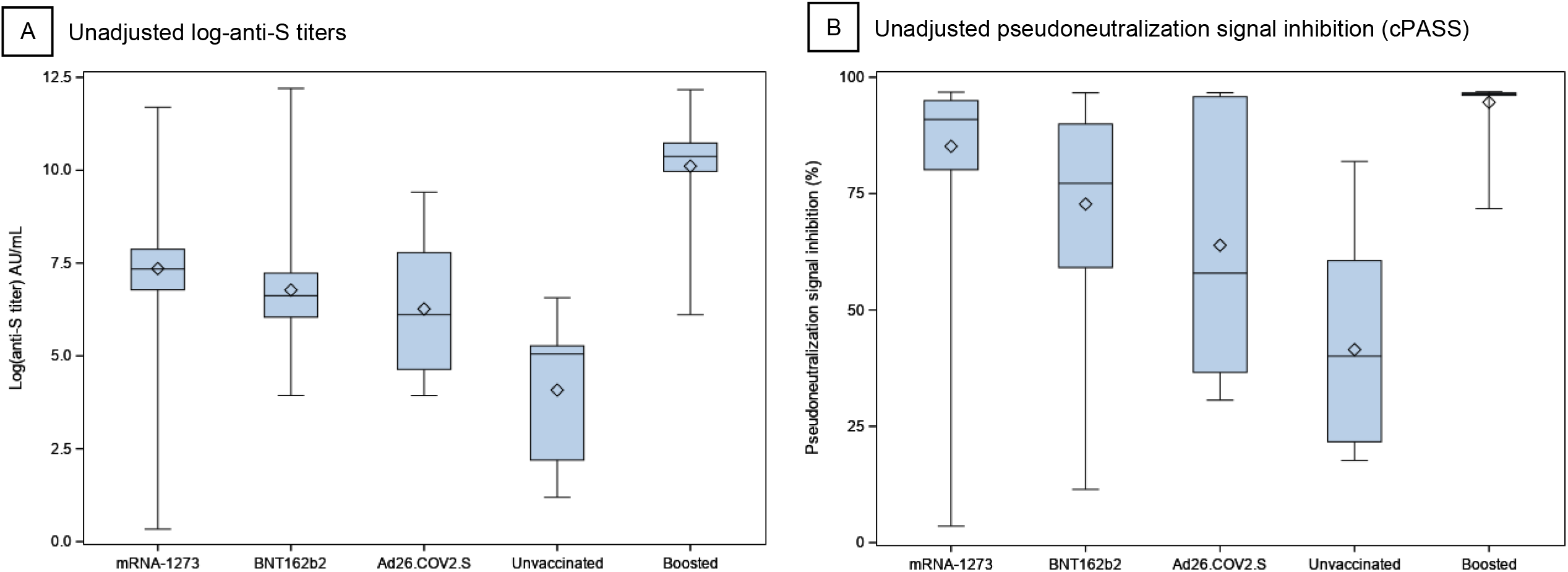
Antibody Distribution by COVID-19 Vaccine Type. Boxplots showing antibody levels by study group at a median of 8.4 months since complete vaccination. The bar inside each box represents the median and the diamond represents the mean antibody level. A, Difference in unadjusted log-anti-S antibodies by vaccine type. B, Difference in unadjusted pseudoneutralization signal inhibition percentages by vaccine type. Note: Sixty percent (6/10) of the unvaccinated HCWs reported a COVID-19 infection prior to antibody testing.

The survey included questions on demographics, medical history (comorbidities predisposing to worse COVID-19 outcome, immunosuppression), COVID-19 infection history (timing of infection and swab testing, symptoms, and severity), and COVID-19 vaccine status (type, primary vaccination timing, and booster type/timing if done). To account for time lapse between survey completion and blood draw, COVID-19 vaccination/infection status was updated upon blood drawing.

Self-reported COVID-19 infection was based on survey response. Laboratory-confirmed infection was based on medical record evidence of a positive PCR or anti-N antibody test. COVID-19-related hospitalizations were confirmed by medical record review. Breakthrough infection rates were calculated from 14 days after primary vaccination through the day of blood drawing. Unvaccinated HCWs were assessed for infection any time from 241 days prior to their blood draw (the median time from vaccination in the overall cohort [255 days] minus 14 days to the day of blood drawing).

Anti-S antibody tests were performed by Bassett Medical Center’s Laboratory using the Abbott AdviseDx SARS-CoV-2 IgG II semi-quantitative assay with dilution if necessary (Abbott Laboratories, Abbott Park, IL). Anti-N antibody tests were also performed by Bassett’s laboratory using the SARS-CoV-2 IgG Abbott Architect assay (Abbott Laboratories, Abbott Park, IL).^40^

Neutralization antibodies to spike protein RBD were measured using ELISA-based pseudoneutralization (competitive antibody blocking) COVID-19 assays (cPASS™, GenScript, Piscataway, NJ) using USA-WA1/2020 strain genetic background and were performed by Biodesix (Boulder, CO). The α-RBD nAb test measures a subset of antibodies that can block the interaction between the RBD on the SARS-CoV-2 spike protein and the human host receptor angiotensin-converting enzyme 2 (ACE2). Methods for the cPASS surrogate virus neutralization test (SVNT) were previously published, validated, and shown to be 100% sensitive and specific when compared to gold standard, plaque reduction neutralization test (PRNT), with qualitative analysis results 100% in agreement.^41-44^ The nAb assay readout was percent signal inhibition by neutralizing antibodies, calculated as the optical density (OD) value of the sample relative to the OD of the negative control subtracted from one.^42^ The cPASS assay has an EUA for qualitative interpretation of results,^41^ but here we prioritize presenting numeric semi-quantitative percent signal inhibition to compare between-group percentages.

Outcome variables were compared by five prespecified characteristics thought to impact immune responses to COVID-19 vaccination: age, number of comorbidities, immunosuppression, time from completion of primary vaccination (days), and prior COVID-19 infection.

### Statistical Analysis

Categorical variables were compared between groups using chi-square. Fisher’s exact test was used with small cell sizes. Continuous variables were compared using one-by-five analysis of variance (ANOVA). For certain continuous variables with skewed distributions, including time from primary vaccination and pseudoneutralization percentages, we were unable to find a suitable transformation of the data to achieve normality. Therefore, data were converted to ranks prior to being analyzed using ANOVA. For anti-S titers, it was found that a natural log transformation resulted in a normal distribution that could be analyzed in the ANOVA model. Post-hoc tests were performed using Scheffe’s method.

A second set of analyses comparing only mRNA-1273 and BNT162b2 groups was completed. In addition to the main effect of vaccine group, these analyses included an additional dimension for the previously mentioned covariates. Multiple linear regression was also performed on the natural log of anti-S titers and the ranks of the pseudoneutralization percentages, including adjustment for covariates. Age, comorbidities, and time since vaccination were modeled as continuous variables.

Spearman’s correlation coefficients were calculated for anti-S levels and pseudoneutralization percentages. VE was estimated by calculating ((infection rate among the unvaccinated - the infection rate among the vaccinated)/infection rate among the unvaccinated))*100.

Missing data on key variables (COVID-19 infection status, vaccination dates, immunosuppression) were either confirmed directly with study subjects or via medical record review. Other missing values were left as missing; imputation of missing values not conducted. Statistical significance was defined as p<.05. All statistical analyses were performed using SAS version 9.4 (Cary, NC).

## Results

### Participants

Mean age was 49.7, 75.3% were female, 93.5% were white, mean number of comorbidities was 0.7/patient, 4.3% had immunosuppression, and median interval from completion of primary vaccination to blood drawing was 255 days (8.4 months). Baseline variables were balanced by age, sex, race, and comorbidities (p>.05), not immunosuppression (p=.047), prior COVID-19 infection (p<.001), time since vaccination (p<.001), and vaccine choice (p<.001) (Table 1).

**Table 1.** Demographics and Clinical Characteristics of Participants by COVID-19 Vaccine Status.

| **Characteristic** | **Total** | **Group 1**<br>**(mRNA-1273)** | **Group 2**<br>**(BNT162b2)** | **Group 3**<br>**(Ad26.COV2.S)** | **Group 4**<br>**(Unvaccinated)** | **Group 5**<br>**(Boosted)** | **P value<sup>a</sup>** |
| --- | --- | --- | --- | --- | --- | --- | --- |
| **No. (%)** | 647 | 387 (59.8) | 212 (32.8) | 10 (1.5) | 10 (1.5) | 28 (4.3) |  |
| **Age, years** |  |  |  |  |  |  |  |
| Mean (SD) | 49.7(12.8) | 50.1 (12.7) | 49.3 (12.6) | 49.7 (10.0) | 40.6 (13.8) | 50.8 (15.4) | .21 |
| <50 | 304 (47.0) | 181 (46.8) | 98 (46.2) | 5 (50) | 8 (80) | 12 (43) | .33 |
| >50 | 343 (53.0) | 206 (53.2) | 114 (53.8) | 5 (50) | 2 (20) | 16 (57) |  |
| **Sex<sup>b</sup>** |  |  |  |  |  |  |  |
| Men | 148 (22.9) | 95 (24.5) | 42 (19.8) | 2 (20) | 0 (0) | 9 (32) | .19 |
| Women | 487 (75.3) | 284 (73.3) | 167 (78.8) | 7 (70) | 10 (100) | 19 (68) |  |
| **Race<sup>b,c</sup>** |  |  |  |  |  |  |  |
| Asian | 10 (1.6) | 7 (1.8) | 2 (0.9) | 0 (0) | 0 (0) | 1 (4) | .51 |
| Black | 6 (0.9) | 4 (1.0) | 2 (0.9) | 0 (0) | 0 (0) | 0 (0) | .99 |
| White | 605 (93.5) | 356 (92.0) | 205 (96.7) | 7 (70) | 10 (100) | 27 (96) | .067 |
| American Indian/Alaskan Native | 3 (0.5) | 2 (0.5) | 0 (0.0) | 1 (10) | 0 (0) | 0 (0) | .054 |
| Other | 11 (1.7) | 6 (1.6) | 4 (1.9) | 1 (10) | 0 (0) | 0 (0) | .35 |
| **Comorbidities** |  |  |  |  |  |  |  |
| Mean # conditions per person (SD) | .7 (1.0) | .8 (1.0) | .7 (1.0) | 1.1 (1.2) | .4 (1.0) | .8 (1.1) | .60 |
| 0-1 | 525 (81.1) | 315 (81.4) | 171 (80.7) | 7 (70) | 9 (90) | 23 (82) | .85 |
| 2+ | 122 (18.9) | 72 (18.5) | 41 (19.3) | 3 (30) | 1 (10) | 5 (18) |  |
| **Immunosuppression<sup>b</sup>** |  |  |  |  |  |  |  |
| Yes | 28 (4.3) | 16 (4.1) | 7 (3.3) | 0 (0) | 0 (0) | 5 (19) | .047 |
| No | 608 (94.0) | 366 (94.6) | 201 (94.8) | 10 (100) | 9 (90) | 22 (82) |  |
| **Ever infected with COVID-19<sup>d</sup>** |  |  |  |  |  |  |  |
| Yes | 57 (8.8) | 32 (8.3) | 15 (7.1) | 3 (30) | 6 (60) | 1 (4) | <.001 |
| No | 590 (91.2) | 355 (91.7) | 197 (92.9) | 7 (70) | 4 (40) | 27 (96) |  |
| **Time since vaccination** |  |  |  |  |  |  |  |
| Median interval between primary vaccination and blood draw (days) | 255.0 | 255.0 | 254.0 | 185.0 | -- | 285.0 | <.001 |
| <8.4 months | 316 (48.8) | 193 (49.9) | 110 (51.9) | 10 (100) | -- | 3 (11) | <.001 |
| >8.4 months | 321 (49.6) | 194 (50.1) | 102 (48.1) | 0 (0) | -- | 25 (89) |  |
| **Chose vaccine type<sup>b,e</sup>** |  |  |  |  |  |  |  |
| Yes | 178 (27.5) | 114 (29.5) | 47 (22.2) | 10 (100) | -- | 7 (25) | <.001 |
| No | 455 (70.3) | 271 (70.0) | 163 (76.9) | 0 (0) | -- | 21 (75) |  |

<sup>a</sup>P value for overall comparison across all groups. X<sup>2</sup> test was used for comparisons of categorical variables and 1-by-5 ANOVA for continuous variables.
<sup>b</sup>Percentage does not add to 100% due to missing data.
<sup>c</sup>Race was self-reported from 5 fixed non-mutually exclusive categories. "Other" race indicates the HCW provided a response different from the five categories. Ethnicity data was excluded due to a low response rate (54%).
<sup>d</sup>Self-reported and/or laboratory-confirmed infection. Self-reported COVID-19 infection was identified by a positive response to the survey question: "To your knowledge, do you have or have you ever had COVID-19?" Laboratory-confirmed infection was based on medical record evidence of a positive PCR test or a positive anti-N antibody test from the original 2020 or this follow-up study.
<sup>e</sup>Response to survey question: "Did you choose your vaccination site/time based on which vaccine was being offered?"

Among the 28 boosted HCWs, 19 (67.9%) received BNT162b2 (primary)-BNT162b2 (booster), 5 (17.9%) mRNA-1273-mRNA-1273 3^rd^ dose, 2 (7.1%) mRNA-1273-mRNA-1273 booster, and 2 (7.1%) mRNA-1273-BNT162b2.

### Anti-S Antibodies

Unadjusted median (IQR) anti-S titers were highest in Group 5 [31898.8 AU/mL (21347.1-45820.1)] and lowest in Group 4 [113.4 AU/mL (3.7-194.0)]; significantly higher in Group 1 [1539.5 AU/mL (876.7-2626.7)] than Group 2 [751.2 AU/mL (422.0-1381.5)]; and not significantly higher in Groups 1 and 2 than Group 3 [451.6 AU/mL (103.0-2396.7)] (overall, p<.001; Groups 1 vs. 2, p<.001; Groups 1, 2, or 5 vs. 4, p<.006; Groups 1, 2, 3, or 4 vs. 5, p<.001; all other comparisons, p>.05) (Table 2/Figure 1). In multivariable analysis, adjusted median anti-S titers for Groups 1 and 2 remained significantly different (p<.001) (Table 3). All covariates were independently associated with anti-S titers except for number of comorbidities (p=.93).

**Table 2.** Unadjusted Antibody Levels by COVID-19 Vaccine Type.

|  | Group 1<br>(mRNA-1273) | Group 2<br>(BNT162b2) | Group 3<br>(Ad26.COV2.S) | Group 4<br>(Unvaccinated) | Group 5<br>(Boosted) | Pairwise *P* values |
| --- | --- | --- | --- | --- | --- | --- |
| **Anti-S antibody titer, AU/mL** |  |  |  |  |  |  |
| No. | 387 | 212 | 10 | 10 | 28 |  |
| Percent positive ($\geq 50$ AU/mL) | 99.5 | 100 | 100 | 60 | 100 | $p < .001$ |
| Median (IQR)<sup>a</sup> | 1539.5<br>(876.7-2626.7) | 751.2<br>(422.0-1381.5) | 451.6<br>(103.0-2396.7) | 113.4<br>(3.7-194.0) | 31898.8<br>(21347.1-45820.1) | Group 1 vs. 2: $p < .001$<br>Group 1 vs. 3: $p = .10$<br>Group 1 vs. 4: $p < .001$<br>Group 2 vs. 3: $p = .99$<br>Group 2 vs. 4: $p = .006$<br>Group 3 vs. 4: $p = .23$<br>Group 5 vs. all: $p < .001$ |
| Age, years |  |  |  |  |  |  |
| <50 | 1668.9 | 989.4 | 915.2 | 82.7 | 32855.0 | Group 1 vs. 2: $p < .001$ |
| $\geq 50$ | 1414.8 | 662.3 | 273.7 | 186.7 | 30684.8 | Group 1 vs. 2: $p < .001$ |
| *p*-value<sup>b</sup> | .40 | .002 |  |  |  |  |
| Comorbidities |  |  |  |  |  |  |
| 0-1 | 1487.4 | 709.3 | 477.5 | 156.4 | 30374.2 | Group 1 vs. 2: $p < .001$ |
| 2+ | 1783.7 | 800.8 | 425.7 | 3.3 | 49840.0 | Group 1 vs. 2: $p < .001$ |
| *p*-value<sup>b</sup> | .74 | .97 |  |  |  |  |
| Immunosuppressed |  |  |  |  |  |  |
| Yes | 2093.7 | 423.1 | -- | -- | 22299.8 | Group 1 vs. 2: $p = .002$ |
| No | 1506.2 | 758.8 | 451.6 | 156.4 | 32491.0 | Group 1 vs. 2: $p < .001$ |
| *p*-value<sup>b</sup> | .71 | .25 |  |  |  |  |
| Time between vaccination and testing |  |  |  |  |  |  |
| <8.4 months | 1672.7 | 869.4 | 451.6 | -- | 22299.8 | Group 1 vs. 2: $p < .001$ |
| $\geq 8.4$ months | 1500.8 | 694.6 | -- | -- | 32411.6 | Group 1 vs. 2: $p < .001$ |
| *p*-value<sup>b</sup> | .073 | .42 |  |  |  |  |
| Ever infected with COVID-19<br>(immunologic and/or clinical) |  |  |  |  |  |  |
| Infected | 3604.4 | 5838.8 | 2653.4 | 184.6 | 192509.0 | Group 1 vs. 2: $p = .98$ |
| Uninfected | 1487.4 | 692.4 | 273.7 | 3.5 | 31385.9 | Group 1 vs. 2: $p < .001$ |
| *p*-value<sup>b</sup> | <.001 | <.001 |  |  |  |  |

**Table 2. Unadjusted Antibody Levels by COVID-19 Vaccine Type (continued)**
| **Pseudoneutralization signal inhibition (cPASS) (%)** |  |  |  |  |  |  |
| --- | --- | --- | --- | --- | --- | --- |
| No. | 387 | 211 | 10 | 10 | 28 |  |
| Percent positive ($\geq 30\%$) | 99.0 | 97.2 | 100 | 60 | 100 | p<.001 |
| Median (IQR)<sup>a</sup> | 90.9 (80.1-95.0) | 77.2 (59.1-89.9) | 57.9 (36.6-95.8) | 40.1 (21.7-60.6) | 96.4 (96.1-96.6) | Group 1 vs. 2: p<.001<br>Group 1 vs. 3: p=.25<br>Group 1 vs. 4: p<.001<br>Group 2 vs. 3: p=.99<br>Group 2 vs. 4: p=.028<br>Group 3 vs. 4: p=.24<br>Group 5 vs. all: p<.001 |
| Age, years |  |  |  |  |  |  |
| <50 | 92.0 | 84.7 | 85.5 | 31.1 | 96.4 | Group 1 vs. 2: p<.001 |
| $\geq 50$ | 90.3 | 68.8 | 45.6 | 53.4 | 96.4 | Group 1 vs. 2: p<.001 |
| p-value<sup>b</sup> | .19 | <.001 |  |  |  |  |
| Comorbidities |  |  |  |  |  |  |
| 0-1 | 90.8 | 77.2 | 61.3 | 40.5 | 96.5 | Group 1 vs. 2: p<.001 |
| 2+ | 92.1 | 77.3 | 45.6 | 22.5 | 96.4 | Group 1 vs. 2: p<.001 |
| p-value<sup>b</sup> | .93 | .91 |  |  |  |  |
| Immunosuppressed |  |  |  |  |  |  |
| Yes | 93.1 | 60.7 | -- | -- | 96.1 | Group 1 vs. 2: p=.004 |
| No | 90.8 | 77.3 | 57.9 | 40.5 | 96.5 | Group 1 vs. 2: p<.001 |
| p-value<sup>b</sup> | .95 | .19 |  |  |  |  |
| Time between vaccination and testing |  |  |  |  |  |  |
| <8.4 months | 92.5 | 77.7 | 57.9 | -- | 96.0 | Group 1 vs. 2: p<.001 |
| $\geq 8.4$ months | 90.6 | 77.1 | -- | -- | 96.5 | Group 1 vs. 2: p<.001 |
| p-value<sup>b</sup> | .10 | .74 |  |  |  |  |
| Ever infected with COVID-19 (immunologic and/or clinical) |  |  |  |  |  |  |
| Infected | 95.8 | 96.0 | 96.5 | 52.1 | 96.7 | Group 1 vs. 2: p>.99 |
| Uninfected | 90.4 | 75.8 | 45.6 | 20.9 | 96.4 | Group 1 vs. 2: p<.001 |
| p-value<sup>b</sup> | <.001 | <.001 |  |  |  |  |

<sup>a</sup>Overall (across all vaccine groups) p<.001
<sup>b</sup>Within-vaccine group comparison across levels of covariates

**Table 3.** Multivariable Linear Regression Model of Anti-S Antibody Levels (log-transformed)

|  | Regression coefficient (95% CI) | *P* value |
| --- | --- | --- |
| Vaccine type |  |  |
| BNT162b2 (vs. mRNA-1273) | -0.645 (-0.819, -0.472) | <.001 |
| Age, per year | -0.012 (-0.019, -0.005) | <.001 |
| Comorbidities, per condition | -0.004 (-0.087, 0.080) | .93 |
| Immunosuppression |  |  |
| Yes | -0.568 (-0.996, -0.140) | .009 |
| No | Reference |  |
| Previous COVID-19 infection |  |  |
| Yes | 1.444 (1.138, 1.750) | <.001 |
| No | Reference |  |
| Time between vaccination and blood draw, per day | -0.008 (-0.011, -0.006) | <.001 |

### Pseudoneutralization Antibodies

Unadjusted median (IQR) pseudoneutralization signal inhibition percentages were highest in Group 5 [96.4% (96.1-96.6)] and lowest in Group 4 [40.1% (21.7-60.6)]; significantly higher in Group 1 [90.9% (80.1-95.0)] than Group 2 [77.2% (59.1-89.9)]; and not significantly higher in Groups 1 and 2 than Group 3 [57.9% (36.6-95.8)] (overall, p<.001; Group 1 vs. 2, p<.001; Groups 1, 2, or 5 vs. 4, p<.028; Groups 1, 2, 3, or 4 vs. 5, p<.001; all other comparisons, p>.05) (Table 2). In multivariable analysis, adjusted median pseudoneutralization percentages for Groups 1 and 2 remained significantly different (p<.001). Time since vaccination and age were significantly associated with lower percentages (p<.001). Previous infection was associated with higher percentages (p<.001). Immunosuppression (p=.51) and comorbidities (p=.63) were not associated with pseudoneutralization.

### COVID-19 Infection Rates and Vaccine Effectiveness Against Infection (Overall)

COVID-19 infection rates as measured by anti-N antibody and/or clinical laboratory-confirmed COVID-19 infection data during this ∼8.4-month period were 3.4% (13/387) in Group 1, 3.8% (8/212) in Group 2, 20% (2/10) in Group 3, 30% (3/10) in Group 4, and 3.6% (1/38) in Group 5 (overall, p=.004; Groups 1, 2, or 5 vs. 4, p<.05; Group 1 vs. 3, p=.05; Group 2 vs. 3, p=.07; all other comparisons, p>.10). All were treated as outpatients (no hospitalizations or deaths). Estimated VE rates were 89% (95% CI 67%-96%) in Group 1, 87% (95% CI 60%-96%) in Group 2, 33% (95% CI 0%-86%) in Group 3, and 88% (95% CI 0%-99%) in Group 5. Most breakthrough infections were in summer/fall 2021, suggesting Delta variant infections (eFigure 2).

### Subgroups

Stratification by age showed different effects by vaccine type on anti-S antibody titers and pseudoneutralization percentages at 8.4 months, with older HCWs having significantly lower responses than younger HCWs for BNT162b2 (p=.002) but not mRNA-1273 (p=.40). Stratification by number of comorbidities, immunosuppression status, and time from primary vaccination did not show significant differences. However, stratification by prior COVID-19 infection status showed significant effects on anti-S antibody titers and pseudoneutralization percentages for both mRNA-1273 and BNT162b2 (p<.001 for all comparisons) (Table 2).

### Correlation Between Antibody Assays

Correlation for anti-S antibody titers and pseudoneutralization percentages in the overall population was high (ρ=.947, p<.001). Correlation coefficients were .926, .934, .903, .867, and .397 for Groups 1 through 5, respectively (all p<.05).

## Discussion

This COVID-19 vaccine comparative immune response durability cohort study showed that medium range antibody responses at a median of 8.4 months after primary vaccination were higher after vaccination with mRNA-1273 than BNT162b2. Ad26.COV2.S antibody responses were lower but not significantly different than with mRNA-1273 and BNT162b2. Specifically, both median IgG spike protein (anti-S) antibody titers and median anti-USA-WA1/2020 strain RBD pseudoneutralization inhibition percentages were significantly higher with mRNA-1273 than BNT162b2 (1539.5 AU/mL vs. 751.2 AU/mL, p<.001; and 90.9% vs. 77.2%, p<.001). Median anti-S and pseudoneutralization antibodies were dramatically higher in boosted HCWs (31898.8 AU/mL, p<.001 for both) and much lower in unvaccinated HCWs (in whom such antibodies were not nil (113.4 AU/mL and 40.1%, respectively, p<.03 for both), suggesting natural infections (“herd immunity”)). Anti-S titers and pseudoneutralization percentages had high between-assay correlation (ρ=.947) as previously reported.^44^ Differences in antibody responses between mRNA-1273 and BNT162b2 persisted in adjusted analyses. Estimated vaccine effectiveness (VE) against COVID-19 infection was similarly high with both mRNA vaccines (87-89%), but not significantly different than Ad26.COV2.S (33%), although conclusions are limited by high statistical uncertainty due to low event rates, small sample sizes for Ad26.COV2.S and boosted groups, and insufficient post-boosting time to assess its VE.

Prespecified subgroup analyses, restricted to unadjusted comparisons between mRNA-1273 and BNT162b2 groups, found no significant differences in anti-S titers or pseudoneutralization percentages by number of comorbidities, immunosuppression status (in contrast with others’ findings for COVID-19 mRNA vaccines),^17, 45^ or time since primary vaccination.

Conversely, stratification by age showed lower anti-S antibody titers and pseudoneutralization percentages in older (≥ 50 years old) vs. younger (< 50 years old) HCWs with BNT162b2 (anti-S: 662.3 vs. 989.4 AU/mL, p=.002; pseudoneutralization: 68.8% vs. 84.7%, p<.001) but not mRNA-1273 (anti-S: 1414.8 vs. 1668.9 AU/mL, p=.40; pseudoneutralization: 90.3% vs. 92.0%, p=.19). Other studies have shown a similar age effect for the two mRNA vaccines^7, 13^ suggesting that primary vaccination with mRNA-1273 rather than BNT162b2 should be prioritized for older individuals. We did not complete subgroup analyses for VE precluding assessment of such an age effect.

Stratification by prior COVID-19 infection status also showed significant effects, with higher anti-S antibody titers and pseudoneutralization percentages in HCWs with previous COVID-19 infection with mRNA-1273 (anti-S: 3604.4 vs. 1487.4 AU/mL, p<.001; pseudoneutralization: 95.8% vs. 90.4%, p<.001) and BNT162b2 (anti-S: 5838.8 vs. 692.4 AU/mL, p<.001; pseudoneutralization: 96.0% vs. 75.8%, p<.001). Notably, the increased antibody responses seen with prior COVID-19 infection were more pronounced with BNT162b2 than mRNA-1273, particularly for anti-S antibodies (8.4-fold vs. 2.4-fold; and 1.27-fold vs. 1.06-fold, respectively). Other studies have also shown higher antibody responses in previously COVID-19 infected vs. naive individuals with both mRNA vaccines.^4, 46, 47^

This is one of the first comparative COVID-19 vaccine immunologic durability studies; our results are consistent with results from two other studies (below).^3, 35^ A synopsis of the current literature shows many more studies reporting antibody and VE results for individual vaccines rather than comparative analyses.

Individual mRNA-1273 antibody studies show high anti-S, anti-S-RBD, and neutralizing antibody titers post-vaccination that were durable for 3-6 months earlier in the pandemic and increased in immunosuppressed adults upon boosting.^12-15^ VE studies show high protection against infection and severe disease after vaccination (95%), but later in the pandemic when Delta predominated, decreased by 36-46% between 7-9 and 12 months.^27, 28^ A prison outbreak study when Delta predominated, found VE of 56.6% against infection (80.5% in those previously infected) and 84.2% against symptomatic infection.^48^

BNT162b2 antibody studies show high antibody titers (same specific antibodies as above) post-vaccination that wane by 2-6 months (more rapidly in immunosuppressed adults), and increase with boosting.^16-24^ VE studies show high protection against infection with durability for 6 months (90-95%) earlier in the pandemic when Alpha and Beta variants predominated, but with waning to as low as 20% later when Beta and Delta variants predominated; VE against severe disease (hospitalization and death) remained high (>90%) throughout the pandemic.^29-31, 34^ VE against infection, severe infection, and mortality in Israel when Delta predominated was much higher with boosting.^49, 50^

Ad26.COV2.S antibody studies show high titers post-vaccination (albeit lower than with both mRNA vaccines) that are relatively durable for 8 months (only 1.8-fold decrease).^25, 26^ VE studies show protection against infection and severe disease of 64-85% after vaccination.^33, 51^

Comparative antibody studies, soon after primary vaccination, show titers that are similar or higher for mRNA-1273 vs. BNT162b2 and higher with either vs. Ad26.COV2.S.^1-7^ One of two comparative antibody durability studies showed marked decreases in anti-S-RBD and nAb titers for mRNA-1273 and BNT162b2 at 6-8 months. In that study, Ad26.COV2.S’s initially lower titers were more durable such that by 6-8 months titers were similar between the three vaccines.^3^ Another study showed about 3-fold higher peak (14-119 days) anti-S and anti-S-RBD titers with mRNA-1273 vs. BNT162b2 with similar 3-fold decreases after 4 months, such that after 4 months, both antibody levels remained 3-fold higher with mRNA-1273.^35^ Likewise, our results show significantly higher anti-S titers and pseudoneutralization percentages at 8.4 months for mRNA-1273 vs. BNT162b2, and higher (but not significantly different) results for mRNA-1273 and BNT162b2 vs. Ad26.COV2.S. However, we cannot make conclusions about relative diminution of these titers as we did not complete sequential antibody assays.

Comparative VE studies show higher protection against infection and hospitalization for mRNA-1273 (90-95%) and BNT162b2 (90%) than for Ad26.COV2.S (70-85%) post-vaccination.^5, 8-11^ Comparative VE durability studies against infection and severe disease at different pandemic times show waning to 80-95% for mRNA-1273, 65-90% by 3-7 months for BNT162b2, and 60-70% for Ad26.COV2.S;^5, 10, 11, 36-38^ the higher ranges were for VE against more severe disease for all three vaccines. In contrast, our study showed equivalently high VE against infection at 8.4 months (87-89%) with mRNA-1273 and BNT162b2 despite significantly higher anti-S titers and pseudoneutralization percentages with mRNA-1273 than BNT162b2; VE was higher but not significantly different with both mRNA vaccines than with Ad26.COV2.S (33%).

Key study limitations included the following: (1) Our study was observational. We addressed predicted confounders by adjusting for covariates likely to impact results; however, unmeasured confounders (e.g., COVID-19 prevention behavior, vaccine preferences, varying infection rates/exposures, and prevalent variants) may have influenced results. Lack of randomization required standardization of the period during which infection rates and VE were quantified. (2) We did not complete peak antibody assays post-primary vaccination precluding comparisons over time. (3) Analyses of Groups 3-5 were limited by small sample sizes. (4) Significant baseline variable group differences included immunosuppression (higher in Group 5, possibly lowering antibody responses in that group), prior COVID-19 infection rates (higher in Groups 3 and 4, possibly increasing antibody responses in those groups), and time from primary vaccination (higher in Groups 1 and 2, possibly lowering antibody responses in those groups). (5) The study design (with addition of the boosted group) and statistical analysis plan (primary analysis focused on subpopulation of surveyed participants) were modified after initiation of enrollment; however, these changes were prespecified prior to seeing primary outcome data. (6) The boosted group included varying primary vaccination-boosting combinations although BNT162b2-BNT162b2 was predominant (67.9%); and insufficient follow-up time to assess boosting’s effects on VE. (7) Antibody inhibition pseudoneutralization assays were completed rather than gold standard PRNT; however, good correlation has been reported for these tests.^44^

## Conclusions

In this COVID-19 vaccines comparative immunology durability cohort study, at a median of 8.4 months after primary vaccination, IgG spike protein antibody titers and pseudoneutralization inhibition percentages against SARS-CoV-2 USA-WA1/2020 were significantly higher with mRNA-1273 than with BNT162b, both of which were not significantly higher than with Ad26.COV2.S. Vaccine effectiveness against infection remained high with both mRNA vaccines (87-89%), also being higher but not significantly different than with Ad26.COV2.S (33%). Due to statistical uncertainty in the small Ad26.COV2.S group, clinically meaningful outcomes differences cannot be excluded. Boosting led to dramatic increases in anti-S and pseudoneutralization responses.

## Supporting information

eFigure 1

eFigure 2

STROBE checklist

## Data Availability

All data produced in the present study are available upon reasonable request to the authors.

## Acknowledgments

We would like to thank the Bassett Research Institute Center for Clinical Research staff (Melissa Huckabone, Martina King, and Catherine Gilmore) and Casie Collins (New York Center for Agricultural Medicine and Health) for completing the blood draws, the Bassett Healthcare Network Clinical Laboratory staff (Brittany Houghton-Depietro, Kyle Thorn, and Marfas Marakankadavu Parambu) for conducting the anti-S and anti-N antibody assays, and the Biodesix lab staff (Sam Imel and Tristan Finch) for performing the cPass assays. This study would not have been possible without their hard work. We would also like to thank Melinda Hasbrouck for providing logistical support, Tim Dupont for coordinating collaborations between the Bassett Research Institute and Biodesix, and Stephen Clark and Henry Weil for assisting in securing funding to support this study.

## Author Contributions

*Concept and design:* Brunner, Freilich, Victory, Lasher, Fink, Gadomski, Shah

*Drafting of the manuscript:* Brunner, Freilich, Victory, Lasher, Fink, Gadomski

*Critical revision of the manuscript for important intellectual content:* Brunner, Freilich, Victory, Lasher, Fink, Gadomski, Shah

*Statistical analysis:* Krupa, Scribani, Jenkins

*Obtained funding:* Freilich, Shah

*Administrative, technical, or material support:* Brunner, Freilich, Victory, Cross, Bush, Shah, Lasher, Fink, Peek

*Supervision:* Brunner, Freilich, Victory, Lasher, Fink, Gadomski

## References

1. Wang Z, Schmidt F, Weisblum Y, et al. mRNA vaccine-elicited antibodies to SARS-CoV-2 and circulating variants. Nature. Apr 2021;592(7855):616–622. doi:10.1038/s41586-021-03324-6

2. Wheeler SE, Shurin GV, Yost M, et al. Differential antibody response to mRNA COVID-19 vaccines in healthy subjects. Microbiol Spectr. Sep 3 2021;9(1):e0034121. doi:10.1128/Spectrum.00341-21

3. Collier A-rY, Yu J, McMahan K, et al. Differential kinetics of immune responses elicited by Covid-19 vaccines. N Engl J Med. 2021;385(21):2010–2012. doi:10.1056/NEJMc2115596

4. Steensels D, Pierlet N, Penders J, Mesotten D, Heylen L. Comparison of SARS-CoV-2 antibody response following vaccination with BNT162b2 and mRNA-1273. JAMA. Oct 19 2021;326(15):1533–1535. doi:10.1001/jama.2021.15125

5. Self WH, Tenforde MW, Rhoads JP, et al. Comparative Effectiveness of Moderna, Pfizer-BioNTech, and Janssen (Johnson & Johnson) Vaccines in Preventing COVID-19 Hospitalizations Among Adults Without Immunocompromising Conditions — United States, March–August 2021. Vol. 70. Sep 24 2021:1337–1343. MMWR Morb Mortal Wkly Rep.

6. Debes AK, Xiao S, Colantuoni E, et al. Association of vaccine type and prior SARS-CoV-2 infection with symptoms and antibody measurements following vaccination among health care workers. JAMA Intern Med. 2021;181(12):1660–1662. doi:10.1001/jamainternmed.2021.4580

7. Richards NE, Keshavarz B, Workman LJ, Nelson MR, Platts-Mills TAE, Wilson JM. Comparison of SARS-CoV-2 antibody response by age among recipients of the BNT162b2 vs the mRNA-1273 vaccine. JAMA Netw Open. Sep 1 2021;4(9):e2124331. doi:10.1001/jamanetworkopen.2021.24331

8. Pilishvili T, Gierke R, Fleming-Dutra KE, et al. Effectiveness of mRNA Covid-19 vaccine among U.S. health care personnel. N Engl J Med. Sep 22 2021;doi:10.1056/NEJMoa2106599

9. Thompson MG, Stenehjem E, Grannis S, et al. Effectiveness of Covid-19 vaccines in ambulatory and inpatient care settings. N Engl J Med. 2021;385(15):1355–1371. doi:10.1056/NEJMoa2110362

10. Tenforde MW, Self WH, Adams K, et al. Association between mRNA vaccination and COVID-19 hospitalization and disease severity. JAMA. 2021;326(20):2043–2054. doi:10.1001/jama.2021.19499

11. Rosenberg ES, Dorabawila V, Easton D, et al. Covid-19 vaccine effectiveness in New York State. N Engl J Med. 2021;doi:10.1056/NEJMoa2116063

12. Doria-Rose N, Suthar MS, Makowski M, et al. Antibody persistence through 6 months after the second dose of mRNA-1273 vaccine for Covid-19. N Engl J Med. Apr 6 2021;384(23):2259–2261. doi:10.1056/NEJMc2103916

13. Widge AT, Rouphael NG, Jackson LA, et al. Durability of responses after SARS-CoV-2 mRNA-1273 vaccination. N Engl J Med. 2020;384(1):80–82. doi:10.1056/NEJMc2032195

14. Hall VG, Ferreira VH, Ku T, et al. Randomized trial of a third dose of mRNA-1273 vaccine in transplant recipients. New Engl J Med. 2021;385(13):1244–1246. doi:10.1056/NEJMc2111462

15. Benotmane I, Gautier G, Perrin P, et al. Antibody response after a third dose of the mRNA-1273 SARS-CoV-2 vaccine in kidney transplant recipients with minimal serologic response to 2 doses. JAMA. 2021;326(11):1063–1065. doi:10.1001/jama.2021.12339

16. Shrotri M, Navaratnam AMD, Nguyen V, et al. Spike-antibody waning after second dose of BNT162b2 or ChAdOx1. Lancet. 2021;doi:10.1016/s0140-6736(21)01642-1

17. Levin EG, Lustig Y, Cohen C, et al. Waning immune humoral response to BNT162b2 Covid-19 vaccine over 6 months. New Engl J Med. 2021;doi:10.1056/NEJMoa2114583

18. Barriere J, Chamorey E, Adjtoutah Z, et al. Impaired immunogenicity of BNT162b2 anti-SARS-CoV-2 vaccine in patients treated for solid tumors. Ann Oncol. Apr 2021;32(8):1053–1055. doi:10.1016%2Fj.annonc.2021.04.019

19. Massarweh A, Eliakim-Raz N, Stemmer A, et al. Evaluation of seropositivity following BNT162b2 messenger RNA vaccination for SARS-CoV-2 in patients undergoing treatment for cancer. JAMA Oncology. 2021;7(8):1133–1140. doi:10.1001/jamaoncol.2021.2155

20. Eliakim-Raz N, Massarweh A, Stemmer A, Stemmer SM. Durability of response to SARS-CoV-2 BNT162b2 vaccination in patients on active anticancer treatment. JAMA Oncol. 2021;7(11):1716–1718. doi:10.1001/jamaoncol.2021.4390

21. Le Bourgeois A, Coste-Burel M, Guillaume T, et al. Safety and antibody response after 1 and 2 doses of BNT162b2 mRNA vaccine in recipients of allogeneic hematopoietic stem cell transplant. JAMA Netw Open. 2021;4(9):e2126344–e2126344. doi:10.1001/jamanetworkopen.2021.26344

22. Boyarsky BJ, Werbel WA, Avery RK, et al. Antibody response to 2-dose SARS-CoV-2 mRNA vaccine series in solid organ transplant recipients. JAMA. 2021;325(21):2204–2206. doi:10.1001/jama.2021.7489

23. Falsey AR, Frenck RW, Walsh EE, et al. SARS-CoV-2 neutralization with BNT162b2 vaccine dose 3. New Engl J Med. 2021;385(17):1627–1629. doi:10.1056/NEJMc2113468

24. Eliakim-Raz N, Leibovici-Weisman Y, Stemmer A, et al. Antibody titers before and after a third dose of the SARS-CoV-2 BNT162b2 vaccine in adults aged ≥60 years. JAMA. 2021;326(21):2203–2204. doi:10.1001/jama.2021.19885

25. Stephenson KE, Le Gars M, Sadoff J, et al. Immunogenicity of the Ad26.COV2.S vaccine for COVID-19. JAMA. Apr 20 2021;325(15):1535–1544. doi:10.1001/jama.2021.3645

26. Barouch DH, Stephenson KE, Sadoff J, et al. Durable humoral and cellular immune responses 8 months after Ad26.COV2.S vaccination. N Engl J Med. Sep 2 2021;385(10):951–953. doi:10.1056/NEJMc2108829

27. Baden LR, El Sahly HM, Essink B, et al. Phase 3 trial of mRNA-1273 during the Delta-variant surge. New Engl J Med. 2021;doi:10.1056/NEJMc2115597

28. Baden LR, El Sahly HM, Essink B, et al. Efficacy and safety of the mRNA-1273 SARS-CoV-2 vaccine. N Engl J Med. 2020;384(5):403–416. doi:10.1056/NEJMoa2035389

29. Polack FP, Thomas SJ, Kitchin N, et al. Safety and efficacy of the BNT162b2 mRNA Covid-19 vaccine. N Engl J Med. 2020;383(27):2603–2615. doi:10.1056/NEJMoa2034577

30. Thomas SJ, Moreira ED, Kitchin N, et al. Safety and efficacy of the BNT162b2 mRNA Covid-19 vaccine through 6 months. New Engl J Med. 2021;385(19):1761–1773. doi:10.1056/NEJMoa2110345

31. Chemaitelly H, Tang P, Hasan MR, et al. Waning of BNT162b2 vaccine protection against SARS-CoV-2 infection in Qatar. New Engl J Med. 2021;doi:10.1056/NEJMoa2114114

32. Sadoff J, Gray G, Vandebosch A, et al. Safety and efficacy of single-dose Ad26.COV2.S vaccine against Covid-19. N Engl J Med. 2021;384(23):2187–2201. doi:10.1056/NEJMoa2101544

33. Corchado-Garcia J, Zemmour D, Hughes T, et al. Analysis of the effectiveness of the Ad26.COV2.S adenoviral vector vaccine for preventing COVID-19. JAMA Netw Open. 2021;4(11):e2132540–e2132540. doi:10.1001/jamanetworkopen.2021.32540

34. Goldberg Y, Mandel M, Bar-On YM, et al. Waning immunity after the BNT162b2 vaccine in Israel. N Engl J Med. Oct 27 2021;doi:10.1056/NEJMoa2114228

35. Bajema KL, Dahl RM, Evener SL, et al. Comparative Effectiveness and Antibody Responses to Moderna and Pfizer-BioNTech COVID-19 Vaccines among Hospitalized Veterans — Five Veterans Affairs Medical Centers, United States, February 1–September 30, 2021. Vol. 70. Dec 10 2021:1700–1705. MMWR Morb Mortal Wkly Rep.

36. Grannis S, Rowley EA, Ong TC, et al. Interim Estimates of COVID-19 Vaccine Effectiveness Against COVID-19–Associated Emergency Department or Urgent Care Clinic Encounters and Hospitalizations Among Adults During SARS-CoV-2 B.1.617.2 (Delta) Variant Predominance — Nine States, June– August 2021. Vol. 70. Sep 17 2021:1291–1293. MMWR Morb Mortal Wkly Rep.

37. Andrews N, Tessier E, Stowe J, et al. Vaccine effectiveness and duration of protection of Comirnaty, Vaxzevria and Spikevax against mild and severe COVID-19 in the UK. Preprint. medRxiv. Sep 21 2021;doi:10.1101/2021.09.15.21263583

38. Dickerman BA, Gerlovin H, Madenci AL, et al. Comparative effectiveness of BNT162b2 and mRNA-1273 Vaccines in U.S. veterans. New Engl J Med. 2021;doi:10.1056/NEJMoa2115463

39. Brunner WM, Hirabayashi L, Krupa NL, et al. Severe acute respiratory coronavirus virus 2 (SARS-CoV-2) IgG results among healthcare workers in a rural upstate New York hospital system. Infect Control Hosp Epidemiol. Oct 26 2020:1–4. doi:10.1017/ice.2020.1296

40. Bryan A, Pepper MH, Wener SL, et al. Performance characteristics of the Abbott Architect SARS-CoV-2 IgG assay and seroprevalence in Boise, Idaho. J Clin Microbiol. Jul 23 2020;58(8):e00941–20. doi: 10.1128/JCM.00941-20

41. cPass SARS-CoV-2 Neutralization Antibody Detection Kit. 2021. November 12. Accessed August 27 2021. https://www.fda.gov/media/143583/download

42. Petrone L, Petruccioli E, Vanini V, et al. A whole blood test to measure SARS-CoV-2-specific response in COVID-19 patients. Clin Microbiol Infect. Feb 2021;27(2):286 e7–286 e13. doi:10.1016/j.cmi.2020.09.051

43. Tan CW, Chia WN, Qin X, et al. A SARS-CoV-2 surrogate virus neutralization test based on antibody-mediated blockage of ACE2–spike protein–protein interaction. Nat Biotechnol. Sept 1 2020;38(9):1073–1078. doi:10.1038/s41587-020-0631-z

44. Taylor SC, Hurst B, Charlton CL, et al. A new SARS-CoV-2 dual-purpose serology test: Highly accurate infection tracing and neutralizing antibody response detection. J Clin Microbiol. 2021;59(4):e02438–20. doi:doi:10.1128/JCM.02438-20

45. Deepak P, Kim W, Paley MA, et al. Effect of immunosuppression on the immunogenicity of mRNA vaccines to SARS-CoV-2: A prospective cohort study. Ann Intern Med. Nov 2021;174(11):1572–1585. doi:10.7326/m21-1757

46. Forgacs D, Jang H, Abreu RB, et al. SARS-CoV-2 mRNA vaccines elicit different responses in immunologically naïve and pre-immune humans. Front Immunol. Sep 27 2021;12(4000) doi:10.3389/fimmu.2021.728021

47. Abu-Raddad LJ, Chemaitelly H, Ayoub HH, et al. Association of prior SARS-CoV-2 infection with risk of breakthrough infection following mRNA vaccination in Qatar. JAMA. 2021;326(19):1930–1939. doi:10.1001/jama.2021.19623

48. Chin ET, Leidner D, Zhang Y, et al. Effectiveness of the mRNA-1273 vaccine during a SARS-CoV-2 Delta outbreak in a prison. New Engl J Med. 2021;385(24):2300–2301. doi:10.1056/NEJMc2114089

49. Arbel R, Hammerman A, Sergienko R, et al. BNT162b2 vaccine booster and mortality due to Covid-19. New Engl J Med. 2021;doi:10.1056/NEJMoa2115624

50. Bar-On YM, Goldberg Y, Mandel M, et al. Protection against Covid-19 by BNT162b2 booster across age groups. New Engl J Med. 2021;doi:10.1056/NEJMoa2115926

51. Sadoff J, Gray G, Vandebosch A, et al. Safety and efficacy of single-dose Ad26.COV2.S vaccine against Covid-19. New Engl J Med. 2021;384(23):2187–2201. doi:10.1056/NEJMoa2101544

