## Supplementary material for "Comparison of Antibody Response Durability of mRNA-1273, BNT162b2, and Ad26.COV2.S SARS-CoV-2 Vaccines in Healthcare Workers": eFigure 1

**eFigure 1. Flow Diagram of HCWs’ Progress through an Observational Study**

### Identification

Assessed for eligibility (n=1,855)

Missing emails (n=49)

Did not complete blood draw (n=250)

- Scheduling/logistics challenges (n=238)
- Declined to participate (n=12)

Completed blood draw and tested (n=653)

Declined to participate (n=4)

Ineligible (n=8)

Consented and completed survey (n=903)

Sent informed consent and survey (n=1,806)

### Enrollment

### Follow-Up

Excluded due to incomplete primary vaccination (n=6)

Included in primary analysis (n=647)

mRNA-1273 (n=387)

BNT162b2 (n=212)

Ad26.COV2.S (n=10)

Unvaccinated (n=10)

Boosted (n=28)

### Analysis
