## Supplementary material for "Comparison of Antibody Response Durability of mRNA-1273, BNT162b2, and Ad26.COV2.S SARS-CoV-2 Vaccines in Healthcare Workers": eFigure 2

**eFigure 2. COVID-19 Infections by Date and Vaccine Type**

|  |  |
| --- | --- |
| 2020-12-27 | 1 |
| 2021-01-12 | 1 |
| 2021-02-06 | 1 |
| 2021-08-27 | 1 |
| 2020-11-29 | 1 |
| 2020-11-15 | 1 |
| 2021-01-18 | 1 |
| 2021-07-27 | 1 |
| 2020-12-03 | 1 |
| 2021-08-22 | 1 |
| 2020-12-16 | 1 |
| 2020-11-10 | 1 |
| 2021-01-01 | 1 |
| 2020-07-15 | 1 |
| 2020-05-20 | 1 |
| 2020-12-01 | 1 |
| 2020-12-02 | 1 |
| 2020-03-28 | 1 |
| 2021-09-05 | 1 |
| 2020-12-18 | 1 |
| 2020-09-25 | 1 |
| 2020-12-15 | 1 |
| 2021-09-01 | 1 |
| 2020-12-01 | 1 |
| 2021-01-11 | 2 |
| 2021-09-05 | 2 |
| 2020-08-15 | 2 |
| 2021-08-13 | 2 |
| 2020-12-15 | 2 |
| 2020-11-30 | 2 |
| 2020-06-01 | 2 |
| 2020-08-17 | 2 |
| 2021-08-23 | 2 |
| 2021-08-10 | 2 |
| 2020-12-14 | 2 |
| 2021-08-05 | 2 |
| 2021-10-03 | 2 |
| 2021-03-06 | 3 |
| 2021-08-02 | 3 |
| 2021-03-29 | 4 |
| 2020-12-17 | 4 |
| 2021-01-14 | 4 |
| 2021-01-04 | 4 |
| 2021-10-21 | 4 |
| 2021-02-23 | 4 |
| 2021-10-06 | 5 |

mRNA-1273 (n=387)

*

BNT162b2 (n=212)

*

Ad26.COV2.S (n=10)

*

Unvaccinated (n=10)

Boosted (n=28)

Delta

Alpha

Dominant variant in NYS^a^

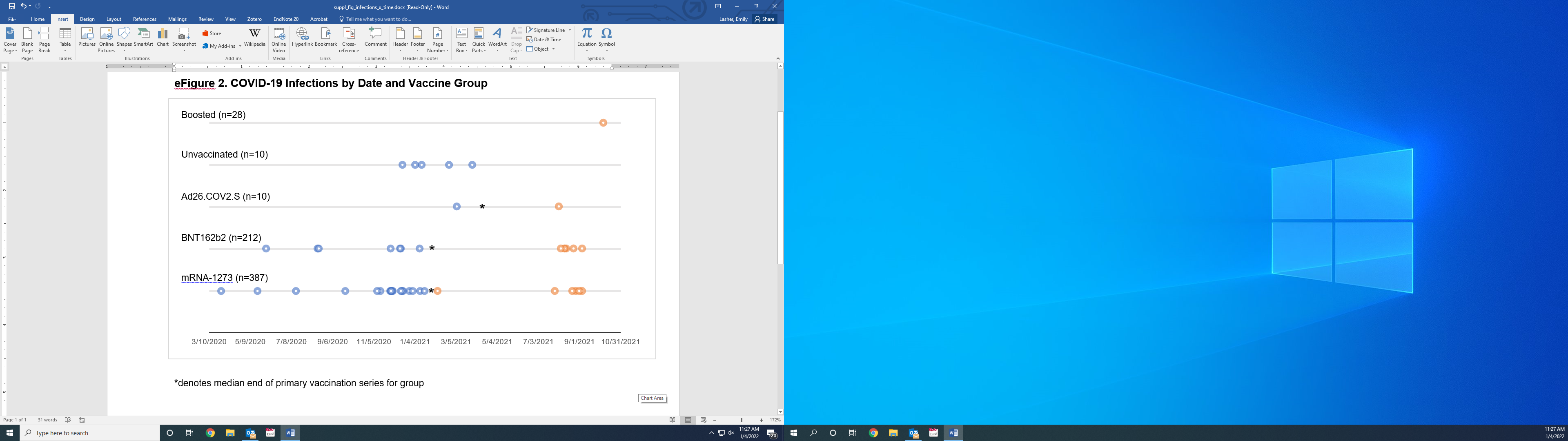
 COVID-19 infection pre-vaccination

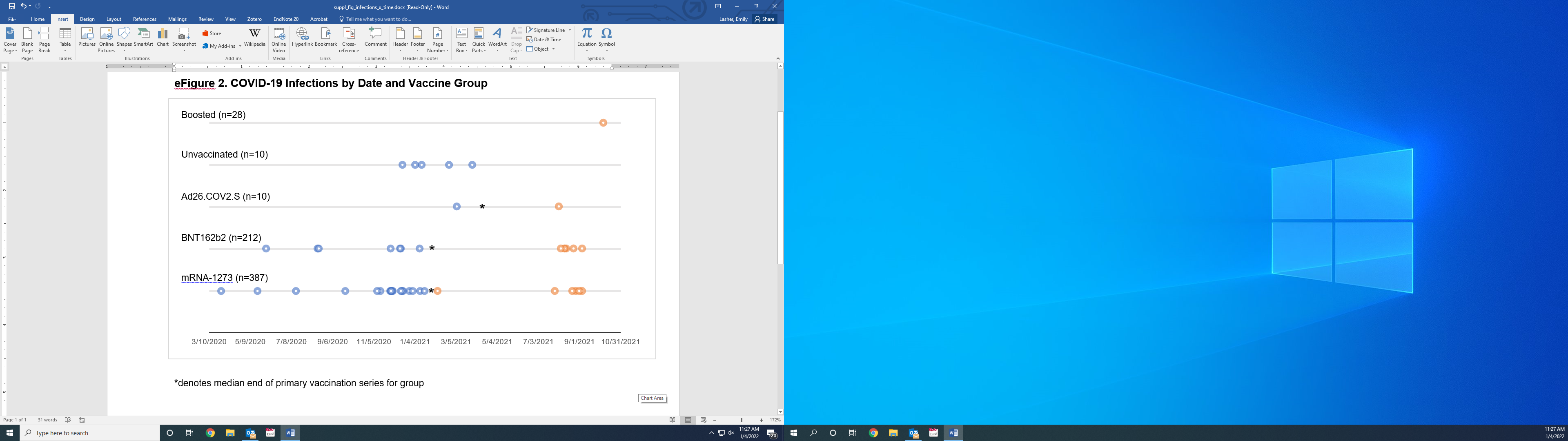
 Breakthrough COVID-19 infection

*

Median end of primary vaccination series

^a^Data from the New York State Department of Health: <https://coronavirus.health.ny.gov/covid-19-variant-data>
